# A Data-Driven Pediatric Type 1 Diabetes Population Health Model Within Specialty Value-Based Care

**DOI:** 10.64898/2026.10.21.26365944

**Authors:** Dominique R. Pahud, Luke A. Harris, Brent Lockee, Katie E. Noland, Jacob Redel, Daniel R. Tilden, Kristy Diederich, Rachel Moran, Mark A. Clements, Juan C. Espinoza

## Abstract

**Objective:** Pediatric specialty value-based care (VBC) for chronic conditions remains under-described. We describe the design and early performance of a data-driven population health model for pediatric type 1 diabetes within a specialty VBC program.

**Research Design and Methods:** Descriptive evaluation at a single academic pediatric medical center and clinically integrated network, 2021–2025, with formal VBC operation beginning in 2024. Participants were children and adolescents with type 1 diabetes attributed through commercial and Medicaid value-based agreements (n=786 in 2025). The program paired infrastructure funding with performance-dependent payments tied to clinical quality and utilization measures, supported by remote monitoring and care management. Measures: percentage receiving the chronic care intervention, population median HbA1c, percentage with HbA1c >9% (75 mmol/mol), percentage with ≥1 diabetic ketoacidosis (DKA) admission, and inpatient and emergency department (ED) visit rates per 1,000/month.

**Results:** From 2021 to 2025, median HbA1c decreased from 8.6% (70 mmol/mol) to 7.9% (63 mmol/mol), HbA1c >9% (75 mmol/mol) decreased from 41.1% to 27.6%, and ≥1 DKA admission decreased from 9.1% to 5.6%. Inpatient admissions per 1,000/month decreased from 11.2 to 7.4 and ED visits from 10.0 to 7.6. The program met 3 of 6 targets in 2024 and all 6 in 2025.

**Conclusions:** Pairing infrastructure funding with performance-dependent payments was feasible and associated with sustained improvements in glycemic control and reduced acute care utilization. Further multi-site evaluation is needed to determine impact, generalizability, and sustainability.

**Graphical Abstract:** 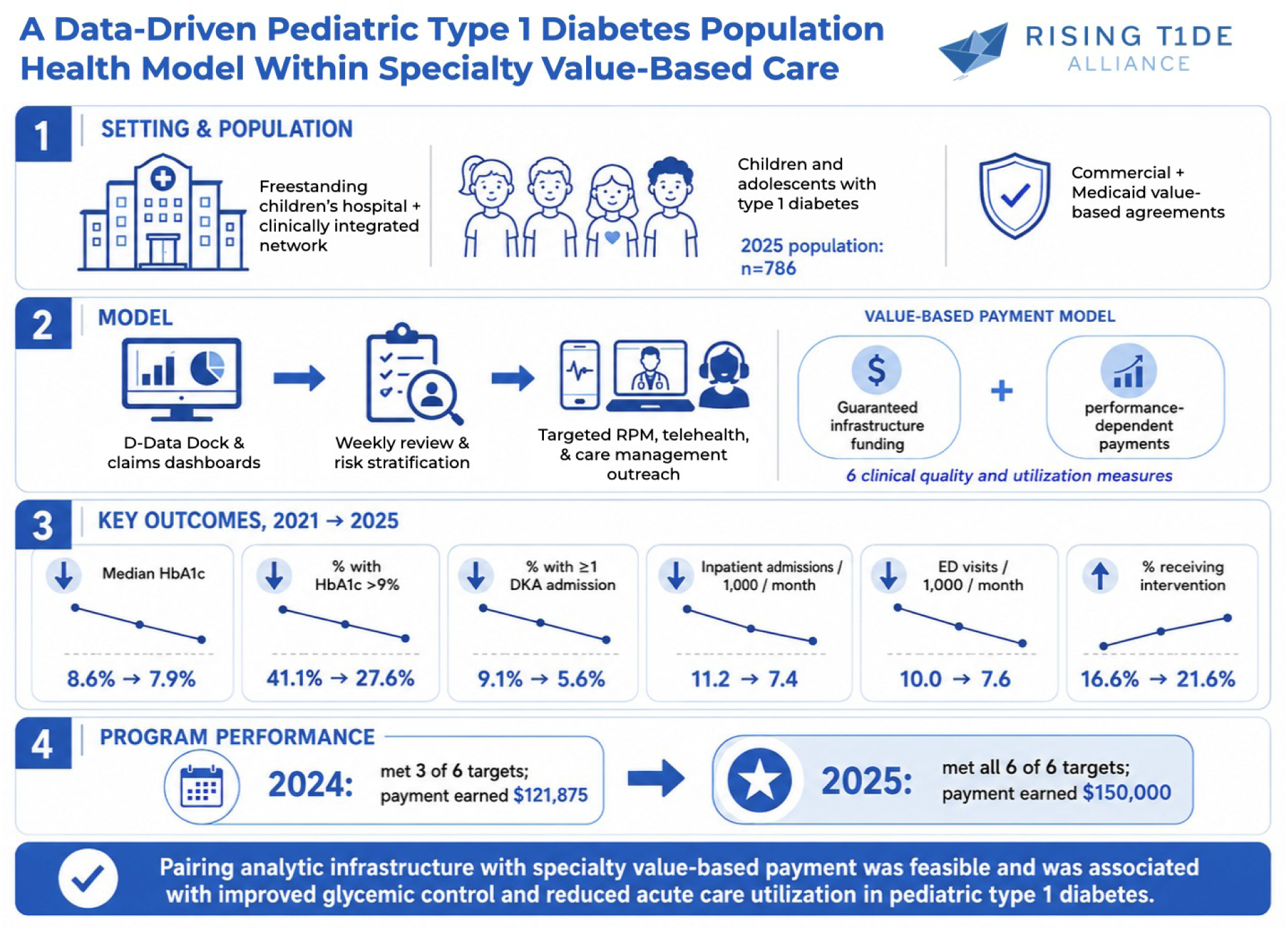

## Introduction

Type 1 diabetes is among the most management-intensive chronic conditions of childhood, requiring continuous insulin delivery, glycemic monitoring, dietary management, and regular multidisciplinary follow-up. Approximately 244,000 children and adolescents younger than 20 years in the United States are living with diabetes, with type 1 diabetes accounting for the majority of pediatric cases.^1^ Contemporary standards of care emphasize team-based management by endocrinology, diabetes education, nutrition, behavioral health, and social work.^2^ Intensive glycemic management reduces the risk of acute complications, particularly diabetic ketoacidosis (DKA), and long-term micro-and macrovascular disease, but achieving and sustaining glycemic targets requires continuous engagement that extends well beyond the clinic visit.^3^

Traditional fee-for-service (FFS) reimbursement remains imperfectly aligned with chronic pediatric care. While some services, such as remote physiologic monitoring, may be reimbursed under existing coding pathways, encounter-based payment still provides limited support for proactive population management, between-visit care coordination, and the technology and workforce investments required for comprehensive chronic care. These constraints are particularly consequential in type 1 diabetes, where achieving glycemic targets depends on continuous engagement between visits (including device data review, insulin dose adjustment, and family education), activities that remain largely unsupported under traditional payment models.^4,5^

Value-based care (VBC) arrangements, which link provider payment to quality and cost outcomes rather than service volume, have demonstrated promise in adult primary care and chronic disease management.^6^ Recent work has explored engaging specialists within accountable care arrangements through nested payment reforms.^7^ However, VBC adoption in pediatric subspecialty care remains limited, in part because many of the most visible alternative payment models have been developed in Medicare populations, whereas pediatric care is financed primarily through Medicaid and commercial insurance, where pediatric-focused value-based arrangements are less mature, less standardized, and often variable across states and plans.^8,9^ Additional challenges include smaller patient cohorts, limited claims-based benchmarking data, complex multidisciplinary care models, and the need for longitudinal engagement spanning years of development. To our knowledge, no published reports describe the design and outcomes of a specialty-level VBC program for pediatric diabetes.

In partnership with Children’s Mercy Integrated Care Solutions (CMICS), the clinically integrated network of Children’s Mercy Kansas City, the Division of Endocrinology and Diabetes launched a chronic condition VBC program for pediatric diabetes in 2024. The program combines guaranteed infrastructure funding with performance-dependent payments tied to 6 clinical quality and utilization measures, creating a financial mechanism to support care model transformation. Pediatric endocrinology was selected as the inaugural specialty for this model based on several criteria: diabetes represents a highly prevalent pediatric chronic condition with established quality metrics; the division had clinical champions comfortable with data-driven quality improvement; and the existing Diabetes Data Dock (D-Data Dock^10^) clinical analytics infrastructure enabled rigorous outcome measurement. This combination positioned the program to demonstrate feasibility while developing replicable operational processes for expansion to other specialty conditions. This descriptive implementation study reports the design, operational infrastructure, and early performance of a data-driven population health model for pediatric type 1 diabetes, delivered within a specialty value-based care program, and draws implications for replication across pediatric subspecialties.

## Research Design and Methods

### Setting and Program Context

Children’s Mercy Kansas City is a freestanding academic pediatric medical center and the region’s primary referral center for pediatric subspecialty care, serving the bi-state Kansas City metropolitan area. The Division of Endocrinology and Diabetes provides specialty care for approximately 2,300 children and adolescents with type 1 diabetes across the region. Children’s Mercy Integrated Care Solutions (CMICS) operates the health system’s clinically integrated network, managing value-based agreements with commercial payers and Medicaid managed care organizations across Missouri and Kansas. In 2024, CMICS established a Chronic Condition Value-Based Care Program, and diabetes was selected as the inaugural specialty condition.

### Program Design and Payment Structure

The VBC program combines guaranteed infrastructure funding with performance-dependent investment opportunity payments. Infrastructure funding supports specialty division investments in care model transformation, including personnel and analytic systems to operationalize a population health care model (see below). Performance-dependent payments are allocated equally between clinical quality (50%) and cost/utilization (50%), with individual measures weighted equally within each domain. Total annual program funding has been $150,000, with the performance-dependent proportion increasing over time from 50% in 2024–2025 to 67% in 2026, with a corresponding decrease in guaranteed infrastructure funding percentage. Payments are directed to the division for infrastructure and care model investments, not individual provider compensation.

### Chronic Care Intervention

The intervention took a population health approach to pediatric type 1 diabetes care, including weekly review of patient data, targeted identification of patients who might benefit from additional support, and care management or telehealth-based outreach as clinically appropriate. Additional detail regarding specific intervention components and related clinical outcomes has been reported previously.^10,11,12,13^ Receipt of chronic care intervention was defined as completion of at least one remote patient monitoring (RPM) encounter with a Certified Diabetes Care and Education Specialist or enrollment in CMICS care management during the measurement year, following identification through the program’s population-level risk-stratification process.

### Performance Measures

Six performance measures were tracked across two domains. Clinical quality measures included (1) percentage receiving chronic care intervention or CMICS care management, (2) population median hemoglobin A1c (HbA1c), (3) percentage with HbA1c greater than 9% (75 mmol/mol), and (4) percentage with 1 or more validated diabetic ketoacidosis (DKA) admissions. Utilization measures included (5) inpatient admission rate per 1,000/month and (6) emergency department visit rate per 1,000/month. Utilization measures were limited to events with a primary diagnosis of diabetes. Measure definitions were applied consistently across all study years.

A validated DKA admission was defined as an inpatient admission with a primary ICD-10 diagnosis code for diabetic ketoacidosis (E08.1x, E09.10, E10.1x, E13.1x), corroborated by laboratory findings within a 24-hour period meeting at least one of the following criteria: blood glucose greater than 200 mg/dL and serum bicarbonate less than 15 mmol/L; blood glucose greater than 200 mg/dL and carbon dioxide less than 15 mmol/L; or blood glucose greater than 200 mg/dL and blood pH less than 7.35. Admissions with a qualifying diagnosis code but insufficient laboratory data to meet any criterion (approximately 10% of coded events) were excluded from the validated count.

### Data Sources and Analytic Pipeline

Clinical and demographic data originated from the institutional electronic health record and were processed through D-Data Dock,^10^ a diabetes-focused analytics platform that generates validated measures. Utilization data were derived from medical claims and eligibility files received by CMICS from participating health plans. CMICS integrated these data into dashboards refreshed monthly. Utilization data for program year 2025 reflect final claims data through December 2025.

### Study Population and Design

This is a descriptive study reporting longitudinal performance trends from 2021 through 2025, with formal program operation beginning in 2024. The study population included children and adolescents with type 1 diabetes attributed to the program through active enrollment in one of CMICS’s value-based agreements during the measurement year, encompassing four Medicaid payers (two in Missouri, two in Kansas) and three commercial payers. The objective was to describe program design, implementation, analytic infrastructure, and early performance trends rather than to estimate a causal effect. Race and ethnicity data were extracted from the electronic medical record as recorded by clinical staff at the time of patient registration.

### Statistical Analysis

Analyses were descriptive. Patient characteristics were summarized using means and standard deviations for continuous variables and counts and percentages for categorical variables. Annual values for each performance measure were calculated and reported descriptively, and longitudinal trends from 2021 through 2025 were summarized in tables and figures. No formal hypothesis testing was performed and no adjusted analyses were conducted. Clinical quality measures were generated by D-Data Dock operating within a Microsoft Azure cloud environment using Databricks for data processing.^10^ Utilization metrics were calculated using Microsoft Power BI. Figures were generated using Claude (claude-sonnet-4-6, Anthropic PBC), as described in the Acknowledgments.

### Ethics Review

This study was reviewed under an exempt protocol approved by the Children’s Mercy Institutional Review Board (STUDY00003967) with a waiver of HIPAA authorization under 45 CFR 164.512(i)(2)(ii). The study met criteria for exemption under 45 CFR 46.104(d) category 4(iii).

### Data and Resource Availability

The datasets generated and analyzed during the current study are not publicly available owing to contractual obligations with participating health plans and the inclusion of protected health information. Aggregate, de-identified data may be available from the corresponding author upon reasonable request. No custom analytic code generated specifically for this study is available; clinical quality measures were produced by the D-Data Dock platform and utilization metrics were calculated in Microsoft Power BI, as described above.

## Results

### Population Characteristics

The VBC-attributed population grew from 661 patients in 2021 to 786 in 2025 (Table 1). Payer mix remained approximately evenly divided between commercial and Medicaid coverage across all years, and demographic characteristics were stable over time, with a mean age of 14.0 (SD, 4.1) years and 48.6% female in 2025. Cohort demographics were broadly consistent with prior national reports of pediatric type 1 diabetes epidemiology.^1,14^

**Table 1.** Population Characteristics by Year.

| Characteristic | 2021 | 2022 | 2023 | 2024* | 2025* |
| --- | --- | --- | --- | --- | --- |
| Total patients, No. | 661 | 735 | 776 | 784 | 786 |
| Payer type, No. (%) |  |  |  |  |  |
| Commercial | 340 (50.1) | 370 (49.8) | 401 (51.2) | 410 (51.7) | 414 (52.0) |
| Medicaid | 339 (49.9) | 373 (50.2) | 382 (48.8) | 383 (48.3) | 382 (48.0) |
| Age, mean (SD), y | 13.7 (3.9) | 13.8 (4.0) | 13.9 (4.1) | 13.9 (4.1) | 14.0 (4.1) |
| Sex, No. (%) |  |  |  |  |  |
| Female | 344 (50.7) | 360 (48.5) | 378 (48.3) | 389 (49.1) | 387 (48.6) |
| Male | 335 (49.3) | 383 (51.5) | 405 (51.7) | 404 (50.9) | 409 (51.4) |
| Race, No. (%) |  |  |  |  |  |
| White | 513 (75.6) | 558 (75.1) | 591 (75.5) | 585 (73.8) | 585 (73.5) |
| Black or African American | 102 (15.0) | 116 (15.6) | 113 (14.4) | 127 (16.0) | 128 (16.1) |
| Hispanic or Latino | 18 (2.7) | 24 (3.2) | 28 (3.6) | 24 (3.0) | 24 (3.0) |
| Other/Unknown† | 46 (6.8) | 45 (6.1) | 51 (6.5) | 57 (7.2) | 59 (7.4) |
| Ethnicity, No. (%) |  |  |  |  |  |
| Hispanic or Latino | 64 (9.4) | 74 (10.0) | 86 (11.0) | 81 (10.2) | 83 (10.4) |
| Not Hispanic or Latino | 610 (89.8) | 663 (89.2) | 689 (88.0) | 707 (89.2) | 707 (88.8) |
| Other/Unknown | 5 (0.7) | 6 (0.8) | 8 (1.0) | 5 (0.6) | 6 (0.8) |

\*VBC program years. †Other/Unknown race includes Asian, Native Hawaiian/Pacific Islander, Multiple Races, American Indian/Alaskan Native, Other, and Unknown. Race and ethnicity reported separately per Office of Management and Budget standards. Percentages reflect patients with complete demographic data.

### Clinical Quality Performance

Clinical quality metrics showed sustained improvement across the observation period (Figure 1). Population median HbA1c decreased from 8.6% (70 mmol/mol) in 2021 to 7.9% (63 mmol/mol) in 2025. The proportion of patients with HbA1c above 9% (75 mmol/mol) decreased from 41.1% to 27.6%, a 33% relative reduction. These improvements were accompanied by an increase in the percentage receiving interventions, from 16.6% to 21.6% over the same period. DKA admissions generally trended downward, with a notable rise to 9.3% in 2024 before declining to 5.6% in 2025.

**Figure 1.**
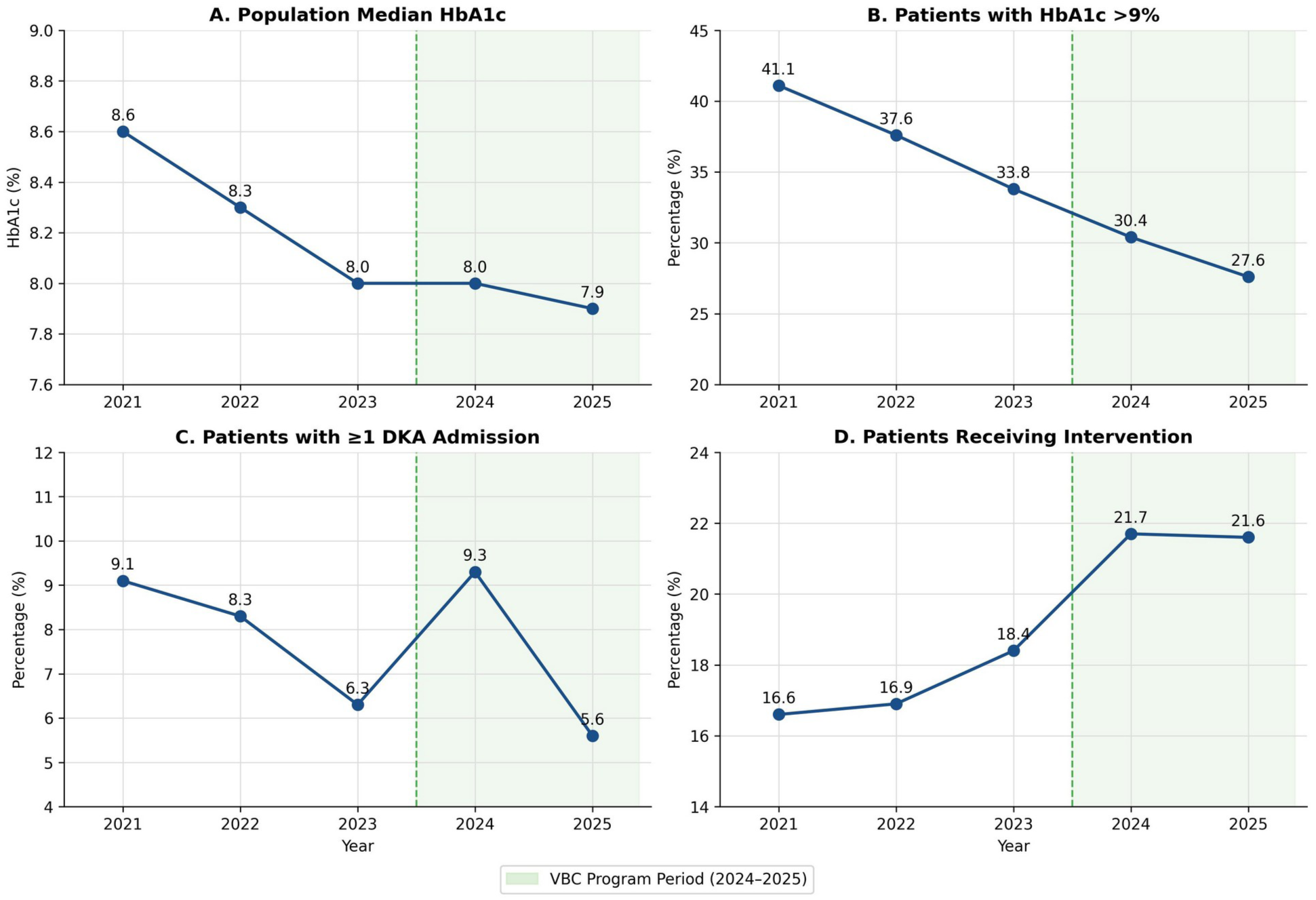
Clinical quality performance trends, 2021–2025. Panels show population median HbA1c, the percentage of patients with HbA1c greater than 9% (75 mmol/mol), the percentage with 1 or more validated diabetic ketoacidosis admissions, and the percentage receiving the chronic care intervention. The shaded area indicates the VBC program period (2024–2025). Source: CMICS dashboard, verified March 2026.

### Utilization Performance

Utilization metrics showed overall improvement (Figure 2). Monthly inpatient admissions per 1,000 decreased from 11.2 in 2021 to 7.4 in 2025, a 34% reduction. Monthly ED visits per 1,000 decreased from 10.0 to 7.6, a 24% reduction.

**Figure 2.**
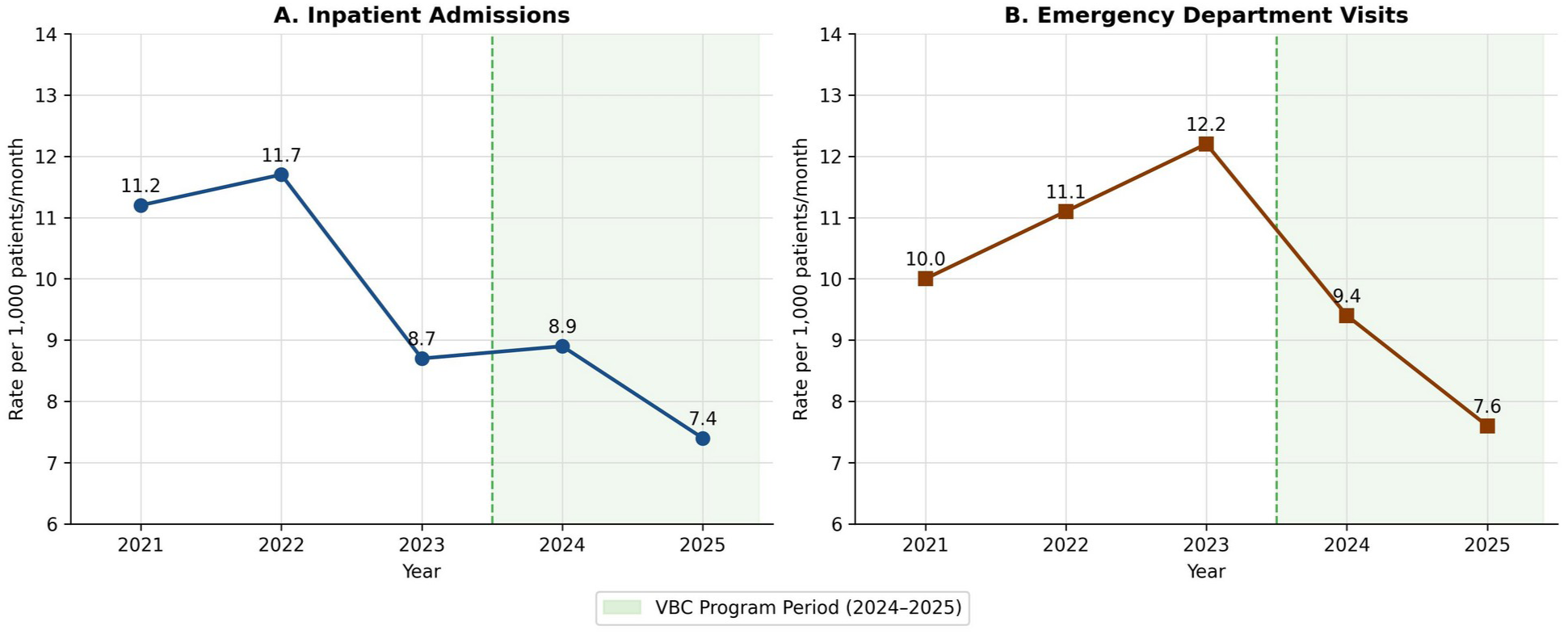
Utilization performance trends, 2021–2025. Panels show inpatient admissions and emergency department visits per 1,000 patients per month, limited to events with a primary diagnosis of diabetes. The shaded area indicates the VBC program period (2024–2025). Source: CMICS dashboard, final CY2025 claims data through December 2025.

### Program Performance

Table 2 summarizes annual performance targets and results. In 2024, the division met 3 of 6 targets (the intervention measure and both utilization measures) and earned $121,875 of the available $150,000. In 2025, performance improved across all measures, and the division met all 6 targets, earning the full $150,000.

**Table 2.** Program Performance Measures, Targets, and Results.

| Measure | 2024 Target | 2024 Actual | Met | 2025 Target | 2025 Actual | Met |
| --- | --- | --- | --- | --- | --- | --- |
| **Clinical Quality** |  |  |  |  |  |  |
| % receiving intervention | 18.6% | 21.7% | Yes | 19.0% | 21.6% | Yes |
| Median HbA1c, % (mmol/mol) | 7.8 (62) | 8.0 (64) | No | 7.9 (63) | 7.9 (63) | Yes |
| % with HbA1c >9% (75 mmol/mol) | 30.0% | 30.4% | No | 29.5% | 27.6% | Yes |
| % with 1+ DKA admission | 6.8% | 9.3% | No | 8.7% | 5.6% | Yes |
| **Cost & Utilization** |  |  |  |  |  |  |
| IP admits per 1,000/mo | 10.4 | 8.9 | Yes | 8.6 | 7.4 | Yes |
| ED visits per 1,000/mo | 15.0 | 9.4 | Yes | 10.0 | 7.6 | Yes |
| **Measures met** |  | **3 of 6** |  |  | **6 of 6** |  |
| **Payment earned** | | **\$121,875** | | | **\$150,000** | |

*Utilization metrics limited to events with a primary diagnosis of diabetes. 2025 utilization data reflect final claims through December 2025.*

## Conclusions

This study describes the design, implementation, and early performance of a data-driven population health model for pediatric type 1 diabetes, delivered within what is, to our knowledge, the first published pediatric specialty value-based care (VBC) program for type 1 diabetes in the United States. In the first two years of the program, we observed improving clinical quality measures accompanied by reduced acute care utilization as more patients received our chronic care intervention. It is worth noting that these were continued or sustained improvement trends from the years preceding the program, with the exception of emergency department visits, which had been increasing in the three years prior. The observed improvements likely reflect multiple concurrent influences, including ongoing quality improvement efforts, broader technology uptake, and evolving standards of pediatric diabetes care. We hypothesize that the VBC program helped support and sustain these trends by providing additional resources and a clear performance framework for population health management, although this study was not designed to evaluate that effect. The program met 3 of 6 performance targets in 2024 and all 6 targets in 2025, resulting in a concomitant increase in performance-based payments. These findings should be interpreted as signals of program performance and feasibility rather than evidence of effectiveness of any one specific intervention.

The increase in DKA admissions observed in 2024 (before returning to prior decreasing trend in 2025) was not attributable to any single identifiable cause through program review. Several institutional changes occurred during this period that, while not causally linked to the observed trend, may have contributed: the acquisition of a community pediatric endocrinology practice, staff turnover within the division, and structural changes to inpatient endocrinology coverage. The absolute number of patients with a validated DKA admission rose from 49 in 2023 to 73 in 2024 before returning to 44 in 2025, a change consistent with small-sample variability in a population of fewer than 800 attributed patients. DKA validation criteria remained unchanged across all study years.

The findings add a practical example to a literature base that has largely emphasized adult populations in accountable care.^6,7^ Published examples of pediatric subspecialty VBC programs remain limited, particularly at the level of a single specialty condition with defined clinical and utilization targets.^8,11^ This report therefore contributes not only early performance data, but also a concrete example of how a pediatric specialty VBC arrangement can be structured, measured, and operationalized within an existing clinically integrated network.

A central feature of the model was its dual-funding structure. Guaranteed infrastructure funding supported upfront investment in the staffing, workflows, and analytic capacity needed for population health management, while performance-dependent payments created accountability for clinical quality and utilization targets. This structure may be particularly relevant in pediatric subspecialty settings, where smaller patient populations and narrower margins can make it difficult for divisions to absorb the costs of care redesign in advance of measurable results.

A further implication of this work is that specialty-level VBC may function not only as a payment mechanism, but also as an organizing structure for population health management. In pediatric settings, value-based arrangements and clinically integrated networks have been used to support care coordination, quality improvement, and provider accountability in ways that extend beyond contracting alone.^11,15^ In this program, the VBC arrangement required explicit measure selection, routine performance review, and shared accountability between the specialty division and the clinically integrated network. These operational features may be as important as the payment model itself, particularly in pediatric subspecialty settings where non-visit-based work is often difficult to measure and sustain.

The selection of pediatric endocrinology as the inaugural specialty was deliberate. Diabetes combined a relatively large attributed population with established quality measures and clinically meaningful utilization outcomes, making it a practical initial condition for specialty-level VBC. Implementation was further supported by the presence of clinical champions, shared governance, and an analytic infrastructure capable of integrating EHR-derived clinical measures with claims-based utilization data, including the D-Data Dock platform.^10^ Following the early success of the diabetes program, CMICS has extended the broader model to epilepsy, juvenile idiopathic arthritis, and cardiology, each time leveraging the framework of the inaugural program. More broadly, this model may offer a starting point for pediatric subspecialties with clearly defined populations, measurable outcomes, and sufficient analytic infrastructure to support population-level management. Collaborative learning networks (e.g., the T1D Exchange^16^) among children’s health systems with value-based payment arrangements represent a natural vehicle for developing pediatric specialty performance benchmarks. As additional institutions implement comparable specialty VBC programs, shared performance data could support the kind of cross-institutional learning that general pediatric population health networks have demonstrated. Where established national specialty collaboratives already track quality metrics, such as those in diabetes, inflammatory bowel disease, and juvenile idiopathic arthritis, their existing benchmarks may also inform performance target-setting for specialty VBC programs, reducing the need to establish new performance standards independently.

Several implementation lessons from the first two program years may inform future pediatric specialty VBC initiatives. Performance target-setting proved iterative and required genuine collaboration between the clinically integrated network and the specialty division. Initial 2024 targets were derived from historical performance trends spanning 2019 through 2023, reviewed jointly at a quarterly Joint Operating Meeting, and finalized through a process in which both teams had standing to propose and contest individual targets. A key structural learning was that setting unified targets across Medicaid and commercial populations required explicit recalibration criteria; the program established a trigger for target review if the Medicaid-to-commercial patient distribution shifted by more than 10 percentage points. Guaranteed infrastructure funding is critical to the program. The ability to direct program payments toward dedicated Certified Diabetes Care and Education Specialist (CDCES) time supporting proactive remote patient monitoring outreach for patients flagged as high-risk was essential to translating population-level risk identification into actionable care. Experience in other settings suggests that risk stratification without the staffing capacity to respond has limited program value, and that performance-only payment structures without an infrastructure component may be insufficient to support the care model transformation required for program success and sustainability. The use of the D-Data Dock^10^ as the program’s analytic infrastructure, a validated clinical data platform generating monthly patient-level quality measures integrated with claims-based utilization data in a shared reporting environment, was a prerequisite rather than an enhancement. This infrastructure made the Division of Endocrinology and Diabetes the natural inaugural specialty for the program, not primarily because diabetes represents a high per-member-per-month cost condition, but because the division had established data capabilities, clinical champions with quantitative fluency, and existing quality improvement infrastructure. Specialty divisions with these characteristics may represent optimal initial partners for pediatric specialty VBC programs, with the operational frameworks and data pipelines developed in an inaugural program providing a foundation extensible to higher-cost specialty conditions. It is important to acknowledge, however, that not all health care organizations have access to comparable analytic infrastructure, staffing, or quality improvement capabilities. A key area for future work will be to identify scalable approaches that lower implementation barriers and enable similar models of high-quality population health management in lower-resource settings.

### Limitations

This study has several limitations. First, as a single-center descriptive study without a control group, it was not designed to estimate a causal effect of the VBC program. Second, improvements observed from 2021 through 2025 may reflect ongoing trends in pediatric diabetes care, including increasing uptake of diabetes technologies and national quality improvement efforts. Third, the study population was limited to patients enrolled in participating CMICS agreements, which may introduce selection effects and limit generalizability. Fourth, this report does not include formal risk adjustment, cost-effectiveness analysis, or patient-level evaluation of which program components were most strongly associated with observed trends.

This descriptive implementation study reports the design, operational structure, and early performance trends of a pediatric specialty VBC program for diabetes. The findings suggest that pairing infrastructure funding with performance-dependent payments is feasible in pediatric type 1 diabetes and can support a measurable population health model for subspecialty care. Further multi-site and comparative evaluation is needed to determine the impact, generalizability, and sustainability of this approach.

## Author Contributions

D.R.P., M.A.C., and L.A.H. led the design and development of the value-based care program. D.R.P. and L.A.H. led overall program development and coordination, including vision-setting, cross-stakeholder coordination across clinical, network, and payer teams, and continuity through launch and early operations. M.A.C. provided founding clinical leadership at the program’s inception, and L.A.H. provided clinically integrated network leadership. M.A.C. led the clinical peer discussion that established the program’s proposed clinical performance targets, with input from B.L. and K.E.N. K.E.N., a certified diabetes care and education specialist, served as the clinical operations lead and led the diabetes educator team that delivered remote patient monitoring engagement to higher-risk patients and families. She also led scheduling coordination for the intervention through most of the program period and directed chart-level review when clinical trends shifted unexpectedly to verify accurate measure characterization. K.D. leads operational oversight and quarterly performance reviews under the direction of L.A.H. J.R. provides current clinical leadership and program oversight, a role previously held by M.A.C. prior to his transition; D.R.T. contributes additional clinical support. B.L. designed and established the clinical data pipelines that produce the program’s monthly clinical quality measures from the institutional electronic health record and oversees their ongoing operation. R.M. manages the utilization and claims data pipelines and conducts the underlying analytic work. Clinical and utilization outputs converge in a shared program dashboard reviewed by both the clinical division and the clinically integrated network. D.R.P. conceived this report, drafted the manuscript, and coordinated co-author review. J.C.E. provided senior-author guidance on framing, scholarly direction, and interpretation, and critically revised the manuscript. D.R.T. contributed to the analytic methods and interpretation of the data. All authors critically reviewed the manuscript, approved the final version, and agree to be accountable for all aspects of the work.

## Statement of Guarantor

J.C.E. is the guarantor of this work and, as such, had full access to all the data in the study and takes responsibility for the integrity of the data and the accuracy of the data analysis.

## Use of Generative Artificial Intelligence

During the course of preparing this work, the authors used Claude (claude-sonnet-4-6, Anthropic PBC) for the purposes of manuscript drafting and editing, table creation and formatting, figure generation, and preparation of submission materials including the cover letter and reporting checklist; and ChatGPT (GPT-5.5, OpenAI) to generate an initial draft of the graphical abstract, which the authors subsequently edited. Following the use of these tools, the authors formally reviewed the content for its accuracy and edited it as necessary. The authors take full responsibility for all the content of this publication.

